# Wastewater-Based Epidemiology for SARS-CoV-2 Biomarkers: Evaluation of Normalization Methods in Small and Large Communities in Southern Germany

**DOI:** 10.1101/2022.07.07.22277349

**Authors:** Alexander Mitranescu, Anna Uchaikina, Anna-Sonia Kau, Claudia Stange, Johannes Ho, Andreas Tiehm, Christian Wurzbacher, Jörg E. Drewes

## Abstract

In the context of the COVID-19 pandemic, wastewater-based epidemiology (WBE) emerged as a useful tool to account for the prevalence of SARS-CoV-2 infections on a population scale. In this study we analyzed wastewater samples from three large (> 300,000 people served) and four small (< 25,000 people served) communities throughout southern Germany from August to December 2021, capturing the fourth infection wave in Germany dominated by the Delta variant (B.1.617.2). As dilution can skew the SARS-CoV-2 biomarker concentrations in wastewater, normalization to wastewater parameters can improve the relationship between SARS-CoV-2 biomarker data and clinical prevalence data. In this study, we investigated the suitability and performance of various normalization parameters. Influent flow data showed strong relationships to precipitation data; accordingly, flow-normalization reacted distinctly to precipitation events. Normalization by surrogate viruses CrAssphage and Pepper Mild Mottle Virus showed varying performance for different sampling sites. The best normalization performance was achieved with a mixed fecal indicator calculated from both surrogate viruses. Analyzing the temporal and spatial variation of normalization parameters proved to be useful to explain normalization performance. Overall, our findings indicate that the performance of surrogate viruses, flow and hydro-chemical data is site-specific. We recommend to test the suitability of normalization parameters individually for specific sewage systems.

**TOC Art:** 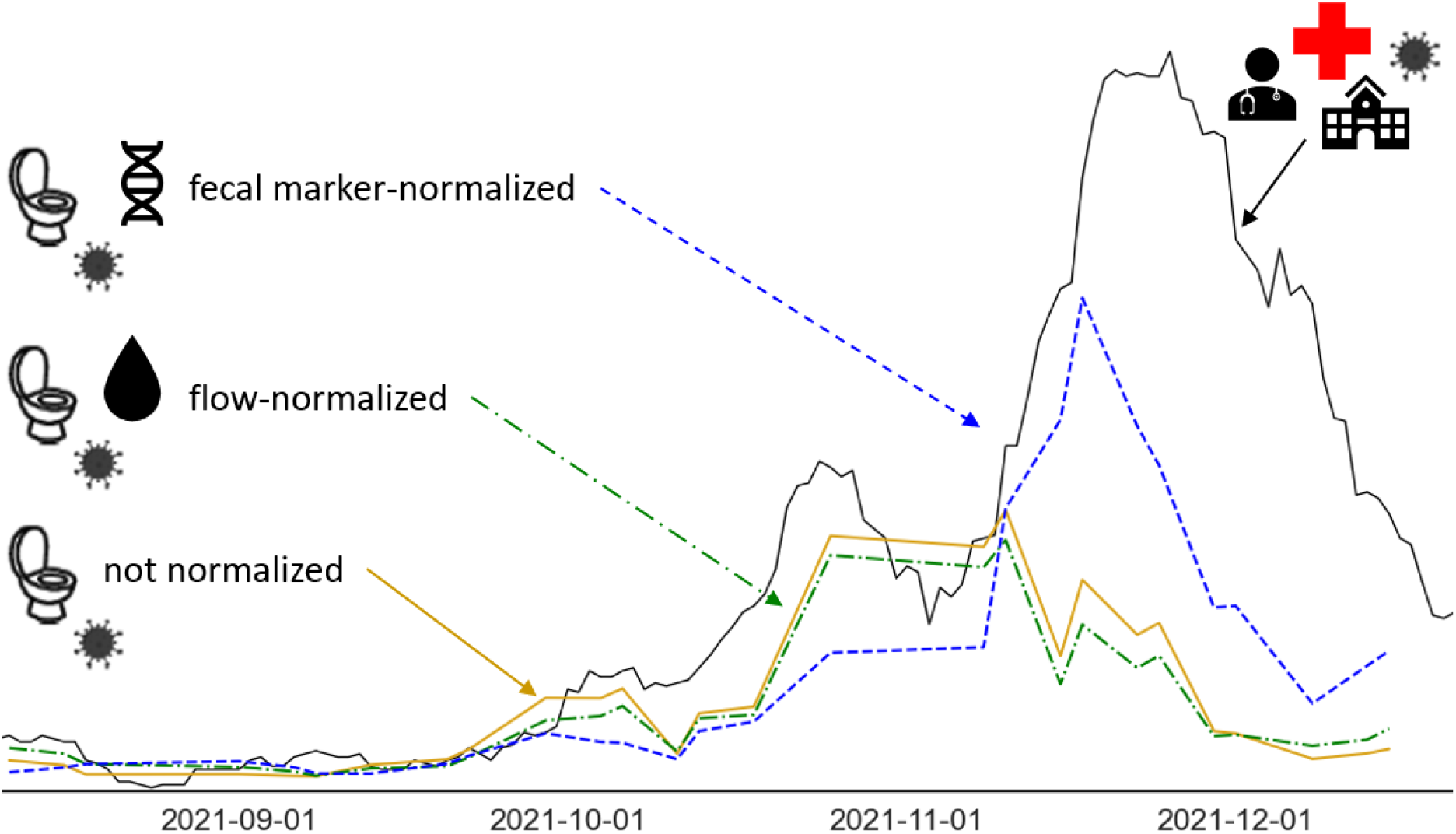

## 1. Introduction

Since late 2019, the COVID-19 pandemic caused by the spread of SARS-CoV-2 impacted public life in large parts of the world. Accounting as precisely as possible for SARS-CoV-2 prevalence in the population has become essential for appropriate public health management. During previous disease outbreaks, wastewater-based epidemiology (WBE) proved to be a useful early warning and monitoring tool for norovirus and poliovirus ^1,2^. Therefore, shortly after the detection of SARS-CoV-2 biomarkers in anal swabs and stool of COVID-19 patients ^3–5^, first WBE studies confirmed the occurrence of SARS-CoV-2 biomarkers in wastewater ^6–9^.

Subsequently, numerous case studies investigated the applicability of SARS-CoV-2 WBE as monitoring and early warning system: from small pilot studies sampling only for several days in the early stage of the pandemic ^10,11^, to long-term ^12,13^ and large-scale ^14^ monitoring programs gathering comprehensive datasets. In the course of the pandemic, knowledge on sampling strategies, quantification methods, data processing, and comparison to epidemiological data increased substantially; yet, there is still intensive ongoing research on various aspects of SARS-CoV-2 WBE.

As the goal of SARS-CoV-2 WBE is to derive valuable information on the infection dynamics in the human population from wastewater, the SARS-CoV-2 biomarker concentrations were frequently compared to epidemiological data. They were observed to strongly correlate with clinical prevalence data and to have lead times of 2 to 24 days ahead of the clinical data, depending on the location and the stage of the pandemic ^7,8,13– 16^, although there are also cases, for which the correlations were low ^17–19^. Furthermore, regression models to predict the viral incidence based on the SARS-CoV-2 biomarker concentrations in wastewater were suggested ^14,16,17^.

However, in order to serve as a suitable method to inform public health, the reliability of the SARS-CoV-2 biomarker concentrations from wastewater has to be assured, as precipitation, industrial and household dischargers, as well as groundwater infiltration do affect wastewater composition and measured SARS-CoV-2 concentrations ^20–22^. Besides data smoothing algorithms ^23^, the main approach to address these dynamic dilution effects in urban wastewater is adjusting SARS-CoV-2 biomarker data with a normalization parameter. For instance, multiplying the gene concentrations with the wastewater flow in order to obtain gene loads has been frequently applied, leading to mixed results ^11,18,24–26^. Furthermore, common wastewater parameters such as ammonia ^14,23^, total nitrogen ^27^, and orthophosphate ^28^ have been used for normalization although they are susceptible to industrial discharge ^29^. Further promising candidates for SARS-CoV-2 biomarker normalization are the pepper mild mottle virus (PMMoV), a plant virus that enters the human body through the diet, and the cross-assembly phage (CrAssphage), a bacteriophage found in the human gut, that are abundant in and closely linked to human feces ^30–32^. Both surrogate viruses have been tested for normalization purposes ^10,12,15,17– 19,24,26,33^. Investigations on the application and comparison of normalization approaches so far focused mainly on flow data, fecal markers CrAssphage and PMMoV, and ammonia ^14,18,19,24,26^. In a further step, several studies assessed the suitability of these parameters for normalization, investigating their temporal and spatial variation ^18,19^, seasonal trends ^34^, or correlation with flow data ^24,26,35^ and precipitation data ^36^.

In this study we report SARS-CoV-2 biomarker data from a wastewater monitoring program of seven wastewater treatment plants (WWTPs) of varying size located in southern Germany. The reported 5-month sampling period captured the fourth wave of infections in Germany, dominated by the Delta variant (B.1.617.2). The aim of this study was to assess different approaches for normalization of SARS-CoV-2 biomarker data (surrogate virus biomarker concentrations, flow data, and electrical conductivity data), combined with a comprehensive analysis (spatial and temporal variation, seasonal trends, relationship to precipitation) of the normalization parameters. We hypothesize that (i) normalization by these water parameters improves the relationship between SARS-CoV-2 wastewater data and clinical prevalence data and that (ii) the comprehensive analysis of the normalization parameters reveals characteristics of the flow situation at the particular sites that can explain the performance of the normalization parameters when applied to SARS-CoV-2 wastewater data.

## 2. Materials and Methods

### 2.1 Sampling Sites, Sample Collection and Transport

Samples were taken from August 2021 to December 2021 at three wastewater treatment plants (WWTP) of large communities (Munich, Augsburg, Karlsruhe) and four WWTPs of smaller communities (Berchtesgaden, Freilassing, Piding, Teisendorf) in the county of Berchtesgadener Land (BGL); all located in southern Germany. The WWTP Berchtesgaden treats the wastewater of several communities, namely Berchtesgaden, Bischofswiesen, Ramsau, and Schönau. The same situation applies to the WWTP Piding treating the wastewater of the communities Anger and Piding. Sampling was performed twice a week except for the WWTP Augsburg from August to November 2021 when it was performed once a week. An overview of the sampling sites and their key characteristics is given in Table 1.

**Table 1:** Overview of sampling sites

| Sampling site | Clinical prevalence data | Precipitation data | Population served | Mean flow rate [$m^3 \cdot d^{-1}$] | Sampling time | Sampling days |
| --- | --- | --- | --- | --- | --- | --- |
| Munich | Robert-Koch Institute (RKI) | CDC, station no. 3379 | 1,156,000 | 278,000 | 24h | 41 |
| Augsburg | Robert-Koch Institute (RKI) | WWTP Augsburg, mean of 11 rain gauges | 343,000 | 144,000 | 24h | 26 |
| Karlsruhe | Robert-Koch Institute (RKI) | CDC, station no. 4177 | 370,000 | 118,000 | 24h | 43 |
| Berchtesgaden | Health Department BGL | CDC, mean of stations no. 364, 515, 516, 6190, 6203 | 21,900 | 10,000 | 3.5h | 40 |
| Freilassing | Health Department BGL | CDC, station no. 1459 | 20,000 | 4,000 | 4h | 42 |
| Piding | Health Department BGL | CDC, mean of stations no. 163, 7424 | 9,000 | 3,500 | 4h | 43 |
| Teisendorf | Health Department BGL | CDC, station no. 5000 | 8,100 | 1,500 | 4h | 43 |

Composite samples were taken at the plant inflow or after the sand trap, using autosamplers with a sampling frequency of 10 minutes. For the large communities, we collected 24 h composite samples since they are less variable than samples of shorter sampling time ^37^. In contrast, for small communities, we collected 3.5-4 h composite samples from 7:00 to 11:00 (resp. 10:30) in order to capture the morning toilet stool peak leading to a temporary high fecal load in wastewater; findings of a recent study corroborate this approach ^38^. The samples were transported to the laboratory at 4 °C.

### 2.2 Sample Processing

Samples from Munich and Augsburg were processed at the Technical University of Munich, Chair of Urban Water Systems Engineering in Garching. Three SARS-CoV-2 specific gene fragments were quantified: the nucleocapsid gene (N2), envelope gene (E), and a sequence from the open reading frame region (ORF). Samples from the WWTP Karlsruhe and the four small communities (Berchtesgaden, Freilassing, Piding, Teisendorf) were processed at the TZW: DVGW-Technologiezentrum Wasser, Department of Water Microbiology in Karlsruhe, Germany. Here, three SARS-CoV-2 specific sequences were quantified: E, ORF and a sequence from the polymerase region (RdRp). The concentrations of biomarkers of the fecal markers PMMoV and CrAssphage for samples from all sites were also determined in this lab according to the same dPCR protocol. See Supplemental Text 1 and Table S1 for further details on sample processing.

### 2.3 Precipitation Data and Clinical Prevalence Data

Precipitation data for the sampling sites Munich, Karlsruhe, Berchtesgaden, Piding, Teisendorf, and Freilassing was obtained from the Climate Data Center (CDC) of the German Meteorological Service ^39^. The weather stations corresponding to the sampling sites are indicated in Table 1. For the sampling site Augsburg, we used precipitation data that was calculated from 11 rain gauges operated by the WWTP Augsburg throughout their service area.

The clinical prevalence data used for the large communities was the publicly available, official data of the 7-days-incidence provided by the Federal Robert-Koch Institute (RKI) in charge of public health surveillance in Germany ^40^. The 7-days-incidence, i.e., the sum of infections of the past seven days in a specific region (e.g., a community) normalized to 100,000 inhabitants, is the prevailing metric used in Germany to measure the infection dynamic. Confirmed SARS-CoV-2 infections were reported daily by the health department of the county BGL for the small communities, attributed to the corresponding sampling sites, and transformed to 7-days-incidences.

### 2.4 Normalization Parameters

The surrogate viruses PMMoV and CrAssphage were available for all sampling sites since they were determined by PCR analysis. Additionally, a mixed fecal indicator (MFI), the mean of the normalized PMMoV and CrAssphage values, was calculated. As PMMoV and CrAssphage abundances do not necessarily have the same order of magnitude, in order to compute MFI, PMMoV and CrAssphage data was normalized to values between 0 and 1 using the maximum value measured for PMMoV and CrAssphage, respectively.

The large WWTPs in Munich, Augsburg, and Karlsruhe provided total daily flows [*m*^3^·*d*^−1^] and daily mean values of the electrical conductivity [μ*S*·*cm*^−1^]. Total daily flows were also provided by the WWTP Berchtesgaden. In contrast, the other small WWTPs do not have a continuous flow measurement and could not provide this information. As a substitute, the mean of the flow [*m*^3^·*s*^−1^] at the beginning and the end of the sampling period was calculated where this flow data was provided (Teisendorf, Piding). The WWTPs in Teisendorf and Berchtesgaden additionally provided measurements of the electrical conductivity during the sampling period. Unfortunately, the WWTP in Freilassing could not provide any additional parameters.

### 2.5 Data Analysis

All data analysis was performed in Python (v3.8.8) using the modules Pandas (v1.4.1), NumPy (v1.20.3), SciPy (v1.7.3), Seaborn (v0.11.2), Matplotlib (v3.3.4), and Statsmodels (v0.12.2).

For the comparison of the spatial variation of the PMMoV and CrAssphage biomarker concentrations, Kruskal-Wallis tests were performed (SciPy v1.7.3) (Fig. 1). The coefficients of variation (CV) for the assessment of the temporal variation of PMMoV, CrAssphage, total daily flows per capita, and electrical conductivity were calculated as the standard deviation divided by the mean (NumPy v1.20.3) (Fig. 1, S1). For details on the linear regression analysis with precipitation as explanatory variable, see Supplemental Text 1.

**Figure 1:**
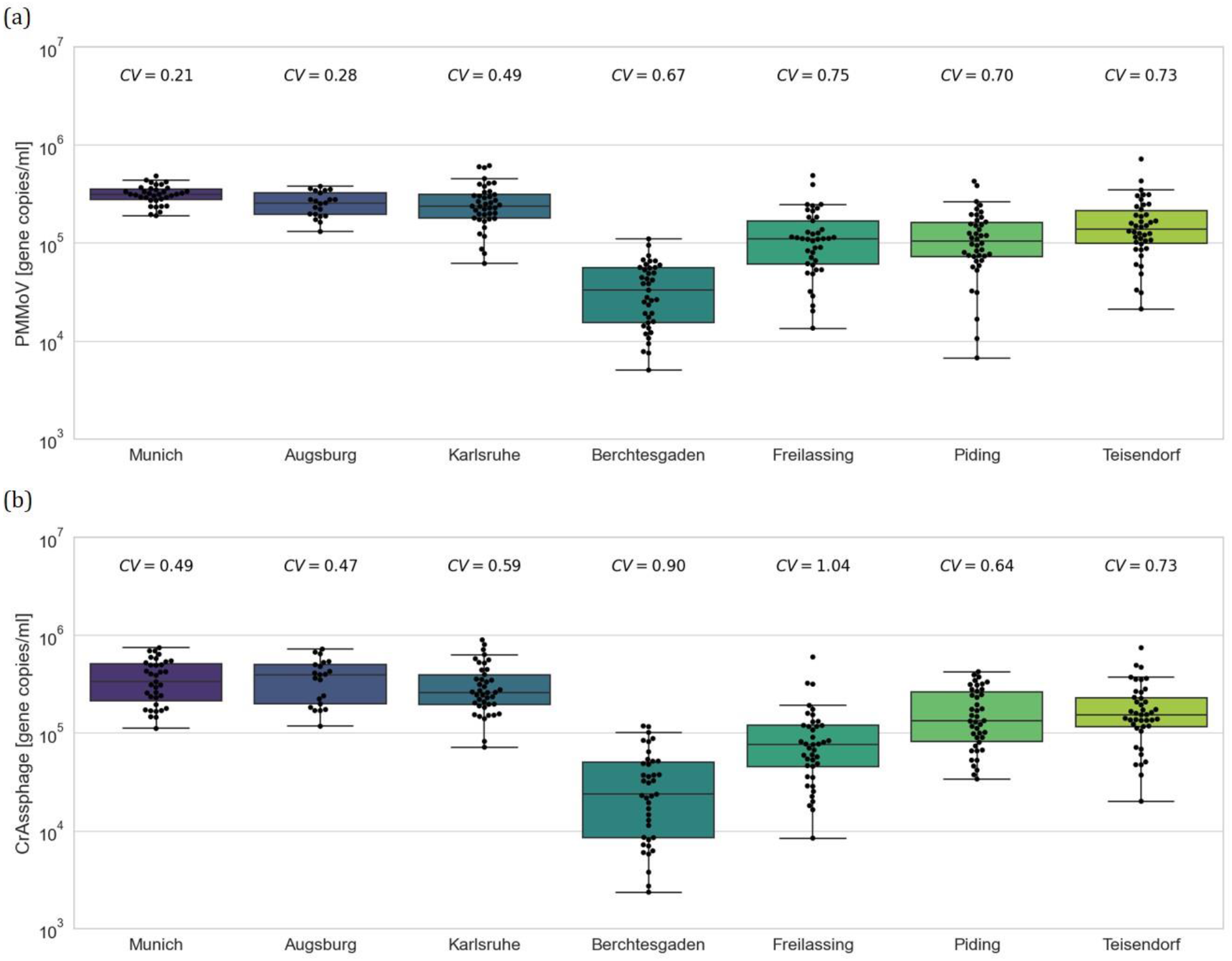
Spatial and temporal variation of PMMoV (a) and CrAssphage (b) for different sampling sites.

After filtering the SARS-CoV-2 biomarker concentrations from the PCR analysis by an automated workflow, the normalizations by the different water parameters were computed (see Supplemental Text 1 for details on automated workflow and application of normalization parameters). Coefficients of determination R^2^ were computed between the SARS-CoV-2 biomarker values and the 7-days-incidence with a linear regression model to assess the performance of the different normalization methods (Fig. 4). The changes in the coefficients of determination R^2^ were classified as “no change” (Δ*R*^2^*≤* ± 0.05), moderate (0.05 < Δ*R*^2^ *≤* 0.2), and considerable (Δ*R*^2^ > 0.2).

## 3. Results and Discussion

### 3.1 Assessment of Normalization Parameters

#### 3.1.1 Analysis of Spatial Variation

When considering the spatial variation of the surrogate viruses among the different sampling sites, we observed that in general, the biomarker concentrations are in a common range from 10^3^ to 10^6^ gene copies/ml (Fig. 1).

More specifically, we observed that the surrogate biomarker concentrations for the large communities Munich, Augsburg and Karlsruhe have similar distributions that are slightly higher than for the small communities Freilassing, Piding, and Teisendorf. Kruskal-Wallis tests confirmed that CrAssphage biomarker distributions for the large communities Munich, Augsburg, and Karlsruhe (*H* = 2.030, *p* = 0.362) and the PMMoV biomarker distributions for Augsburg and Karlsruhe (*H* = 0.053, *p* = 0.818) are not significantly different. Kruskal-Wallis tests additionally showed no significant differences between CrAssphage biomarker distributions for Piding and Teisendorf (*H* = 0.565, *p* = 0.452) and PMMoV biomarker distributions for Piding and Freilassing (*H* = 0.002, *p* = 0.964). Interestingly, the PMMoV and CrAssphage biomarker concentrations for Berchtesgaden were considerably lower than for the other sampling sites.

In previous studies, PMMoV and CrAssphage biomarker concentrations in raw sewage were reported in a similar range to our study ^24,26,34^. The trend of higher biomarker values in large communities we observed was also confirmed by two studies ^10,41^. In contrast, Wilder et al. ^35^ observed an association between larger CrAssphage concentrations and smaller communities. The data of most previous studies, however, did not show notable differences in surrogate virus concentrations of small and large communities ^19,24,26^. We suppose that the particularly low biomarker concentrations for the sampling site Berchtesgaden are a consequence of the relative high groundwater infiltration of up to 54% of the influent flow in this service area located in an alpine region.

As for the total daily flows per capita, values observed for Berchtesgaden and Augsburg were higher than for Munich and Karlsruhe (Fig. S1). This mirrors the ratio of flow and population of the WWTPs’ capacities (Table 1). The elevated values for Augsburg and Berchtesgaden can be explained by a high industrial wastewater intake of approximately 25% and a high groundwater infiltration of up to 54%, respectively. Concerning electrical conductivity, a low spatial variation between sampling sites was generally observed with the exception of Teisendorf, where larger values were stated (Fig. S1). An explanation for these elevated values is that up to 40% of the WWTP’s inflow originates from a local brewery. In literature, brewery wastewater was reported to exhibit high levels of electrical conductivity between 2,440 and 4,710 μ*S*/*cm* ^42^, substantially higher than in urban wastewater.

#### 3.1.2 Analysis of General Temporal Variation and Seasonal Trends

As for the general temporal variation of normalization parameters, we observed that the CV for all parameters examined was lower than 1 except for the CrAssphage data at the sampling site Freilassing (*CV* = 1.04) (Fig. 1, S1). The CV values for PMMoV and CrAssphage are in the same range as the results reported by Greenwald et al. ^19^. Comparing the two surrogate viruses, CrAssphage abundances showed generally higher CV values than PMMoV abundances (Fig. 1). In contrast to these results, Ahmed et al. ^37^ observed a larger temporal variation for PMMoV concentrations than for CrAssphage, whereas Greenwald et al. ^19^ did not observe a clear trend with both PMMoV and CrAssphage showing large and small CV values, depending on the sampling site.

Furthermore, for CrAssphage, PMMoV and electrical conductivity, the small communities overall exhibited a considerably larger CV value than the large communities. This is corroborated by the findings of Holm et al. ^41^ who reported a higher variability of biomarker abundances in small communities. We suppose that in our study, this difference can be explained by the sampling strategy as we performed 24 h composite sampling for large communities and 3.5-4 h composite sampling for small communities in order to cover the morning toilet stool peak but also the size of the sewershed as smaller communities in general show higher variability in quality ^37,43^.

The second aspect of temporal variation investigated was the seasonal trend of the parameters. In our data, for PMMoV and CrAssphage, there were no distinct seasonal fluctuations observable, which is in accordance with observations reported in literature ^31,44^. In contrast, daily flow showed a moderate decrease over the monitoring period for the three sampling sites Augsburg, Munich, and Berchtesgaden, as can be observed best when considering the daily flow values on dry days (Fig. 2, S2). This trend can be explained by the decrease of groundwater levels during a fall season with generally low-precipitation pattern following a summer with high rainfall as it was the case in 2021 in southern Bavaria. Interestingly, this trend was not observed for the sampling site Karlsruhe, where daily flow showed a moderate increase in the same time period (Fig. S2), probably reflecting the disparity between the regional climates of southern Bavaria and the Upper Rhine Plain.

**Figure 2:**
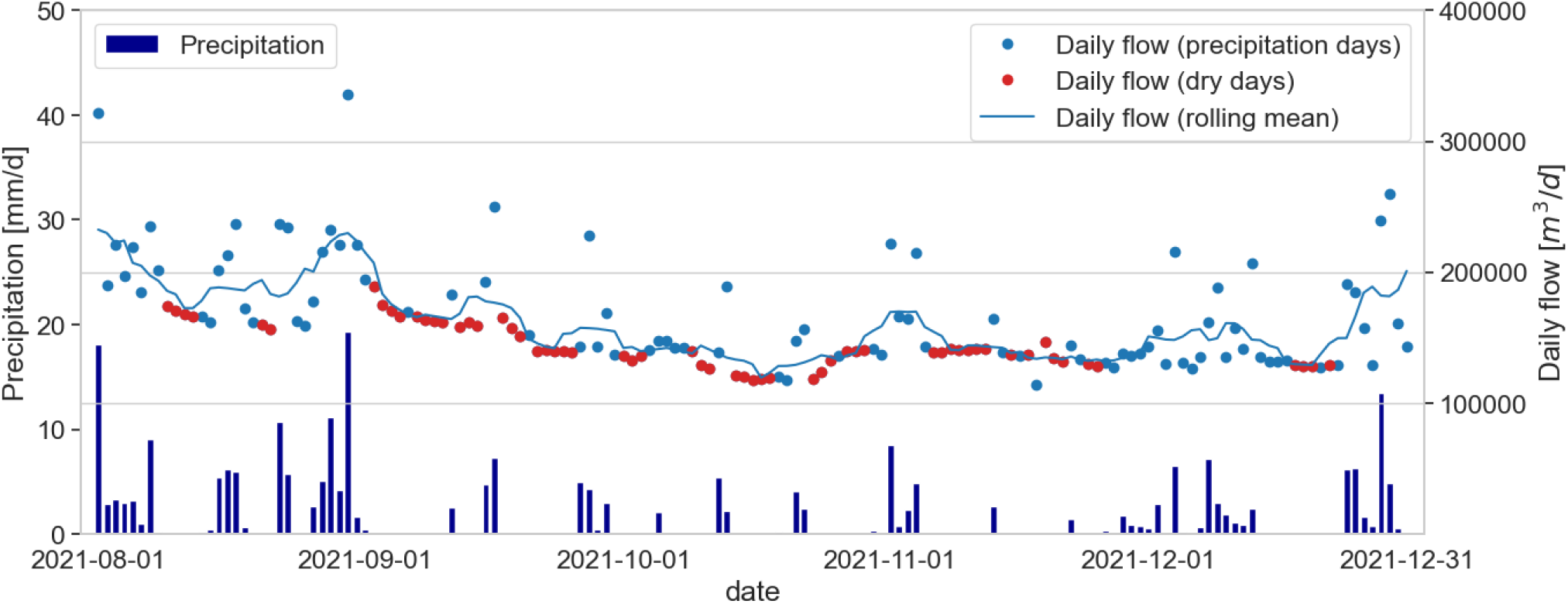
Timeline of daily flow data and precipitation data at sampling site Augsburg.

Additionally, an increase of daily flow values during precipitation events was observed (Fig. 2 and S2), indicating a strong relationship between the two parameters. Linear regression analysis confirmed these findings (Fig. S4). This strong relationship can be explained by the fact that the sewersheds of the sampling sites studied are primarily characterized by combined sewer systems and precipitation events therefore have a major effect on flow volumes. These findings suggest that flow data mirrors precipitation data and thus should be a good normalization parameter to account for dilution effects.

Electrical conductivity showed globally steady levels for all sampling sites except Karlsruhe until the end of November 2021 with lower concentrations during precipitation events (Fig. S3). This reduction during and after precipitation events was considerably more pronounced for the sampling site Karlsruhe (Fig. S3). For Augsburg, Munich, and Berchtesgaden, large variations of the electrical conductivity, including the increase during precipitation periods, were observed in December. These are possibly a consequence of the use of de-icing salt in the snow period that is washed to the sewer system by precipitation or snowmelt. Therefore, the use of electrical conductivity as a normalization parameter should be evaluated carefully because of its possible large variations during the de-icing period. Interestingly, an increase of electrical conductivity in December was not observed for the sampling site Karlsruhe, possibly reflecting the milder regional climate at this sampling site (no snowfall in December 2021) that made the use of de-icing salt unnecessary in this period (Fig. S3).

### 3.2 Application of Normalization Parameters

#### 3.2.1 Analysis of Response of Flow-normalized SARS-CoV-2 Biomarker data to Precipitation Events

From the preliminary assessment of normalization parameters, flow values emerged as a suitable parameter to account for precipitation-driven dilution effects. Examining the late summer and early fall at the sampling site Augsburg, we observed that flow-normalization reacted distinctly to major precipitation events (Fig. 3). In detail, we observed that for abundant and long precipitation periods, e.g. in Augsburg at the end of August 2021, normalizing by flow corrects the low, presumably heavily diluted SARS-CoV-2 biomarker value on 2021-08-30. This flow-normalization is crucial to properly identify an increase in the SARS-CoV-2 biomarker values indicating an acceleration of the infection activity that was confirmed slightly later by the clinical data. In contrast, SARS-CoV-2 biomarker values during dry or low-precipitation periods, e.g. the value in Augsburg on 2021-09-20, are barely changed, just as one would expect. Similar results can be observed for the sampling sites Munich and Karlsruhe (Fig. S5).

**Figure 3:**
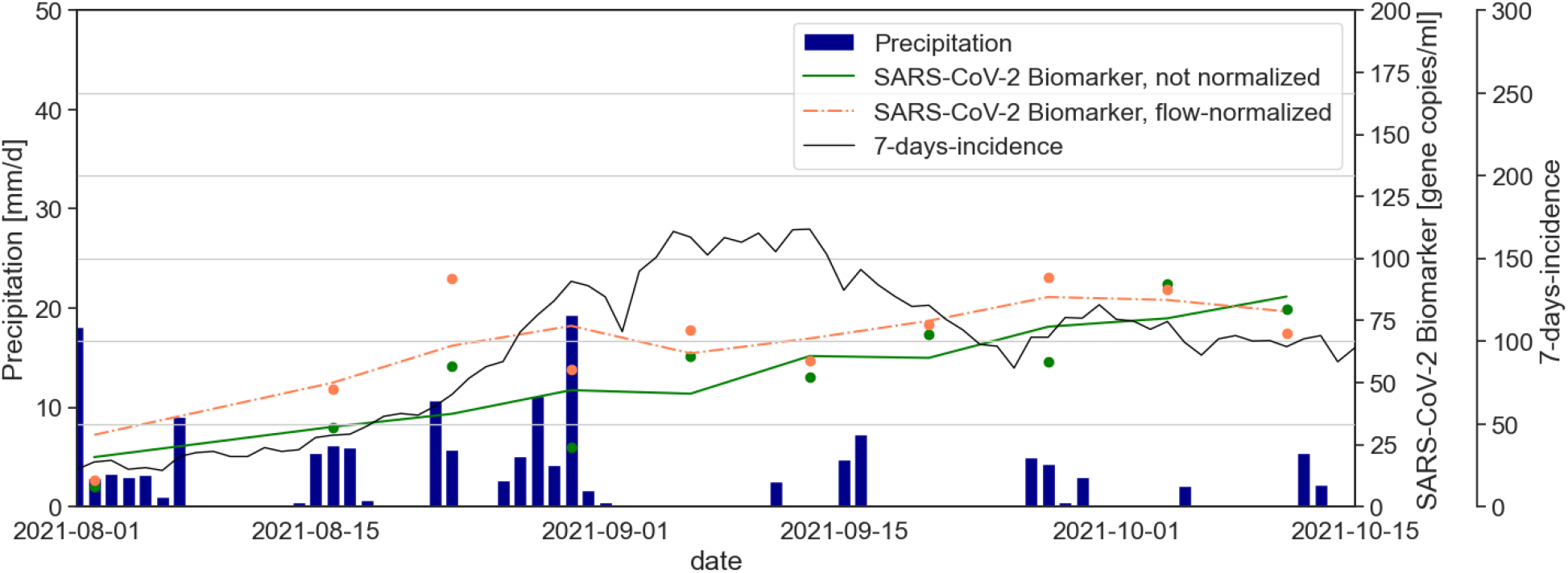
Timeline of precipitation data, SARS-CoV-2 biomarker data (unnormalized and flow-normalized) and 7-days-incidence for sampling site Augsburg in late summer and early fall of 2021.

As total daily flows were not available for the small sampling sites Teisendorf and Piding, a normalization by the mean of the flow at the beginning and the end of the sampling period was applied. For these flow-normalizations, we also observed a reaction to precipitation events although not as strong as for normalizations to total daily flow data (Fig. S5). These results suggest that, in the absence of total daily flow data, normalizing by mean values of the flow during the sampling period can help accounting for precipitation-driven dilution effects.

#### 3.2.2 Analysis of Performance of Different Normalization Methods

In order to compare the performance of all normalization parameters at all sites, linear regression models expressing the relationship between clinical data and the SARS-CoV-2 biomarker concentrations with different normalizations were constructed and the R^2^ values determined (Fig. 4). We observed that generally, R^2^ values are already high (*R*^2^ ≥ 0.7) or moderate (*R*^2^ ≥ 0.4) for most sampling sites without the application of a normalization parameter. Although flow data showed a strong relationship with precipitation data (Fig. 2 and S4) and flow-normalized SARS-CoV-2 biomarker concentrations reacted to precipitation events (Fig. 3), the coefficients of determination for flow-normalized data do not differ substantially from the ones for unnormalized data. R^2^ values are slightly increased for some sites and slightly decreased for others (Δ*R*^2^ *≤* ± 0.05). This is in agreement with recent publications, where correlation coefficients were slightly increased or decreased by flow-normalization compared to unnormalized data, depending on the sampling site (Difference in Spearman’s ρ: ±0.08 ^24^; Difference in Pearson’s r: ±0.11 ^26^). A similar behavior was observed for normalization by electrical conductivity with the exception of the sampling site Karlsruhe where it led to a considerable increase (*R*^2^ = 0.75). This site-specific performance of the parameter electrical conductivity can possibly be explained by the above-mentioned high sensitivity to dilution and by the stable seasonal trend, indicating no distortion by the use of de-icing salt (Fig. S3).

**Figure 4:**
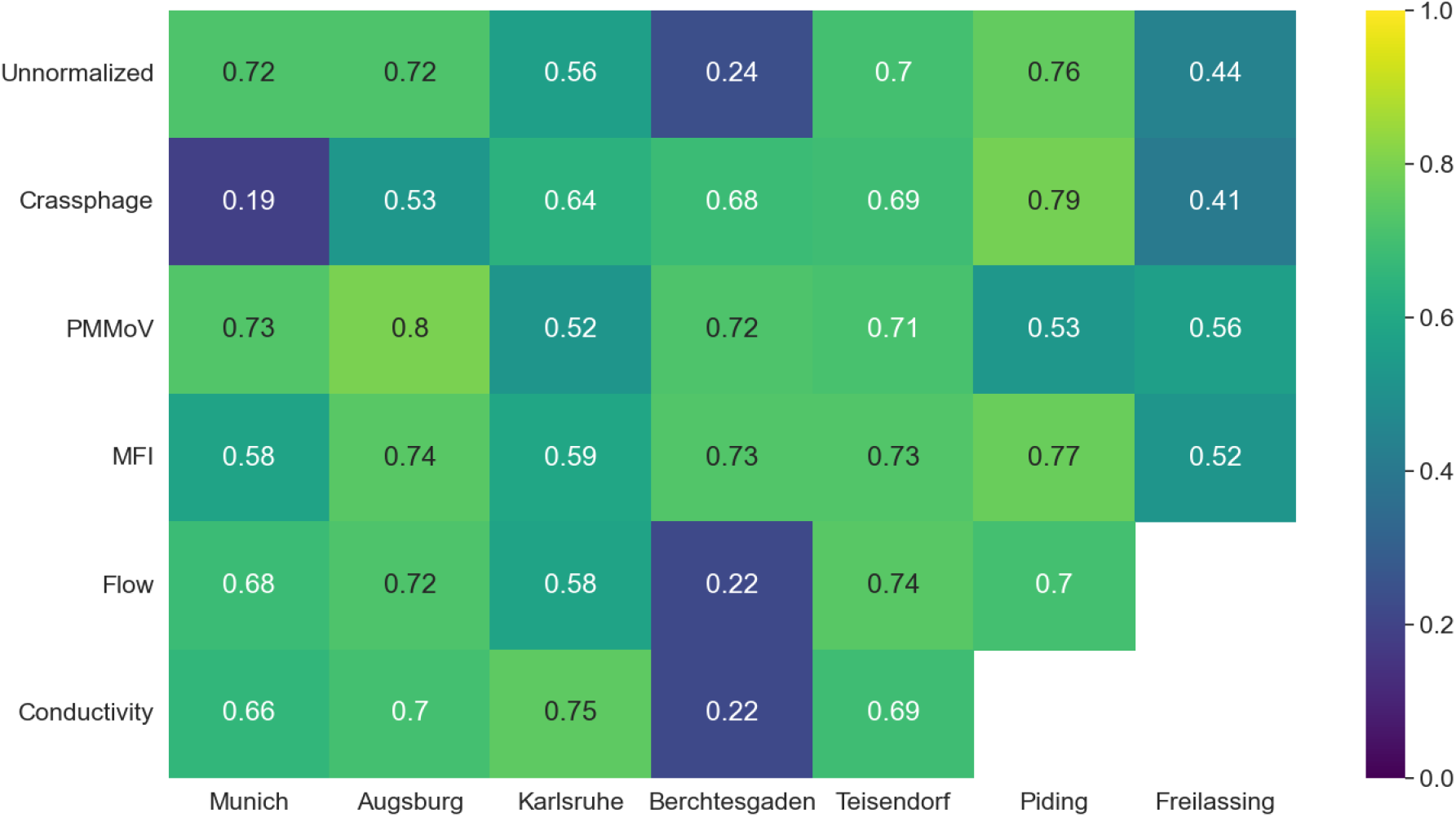
R^2^ of linear regression for clinical prevalence data and SARS-CoV-2 biomarker data for various normalization parameters and sampling sites.

When compared to normalization by surrogate viruses, flow-normalization overall leads to weaker relationships between clinical prevalence data and SARS-CoV-2 biomarker data (Fig. 4). Similar results were obtained in literature, where PMMoV-normalization was observed to lead to stronger correlations with clinical case data than flow-normalization (Difference in Pearson’s r: 0.03 − 0.11 ^26^) or normalization by mass flux through a WWTP (Difference in Pearson’s r: 0.11 − 0.91 ^18^). In contrast, Feng et al. ^24^ reported lower correlation coefficients for PMMoV-normalized data compared to flow-normalized data (Difference in Spearman’s ρ: 0.01 − 0.36). This higher overall normalization performance of surrogate viruses compared to flow can be explained by surrogate viruses not only accounting for stormwater dilution but also for the dynamic variations of fecal load in the wastewater.

The application of surrogate viruses as normalization parameters generally either did not change the R^2^ value (Δ*R*^2^ *≤* ± 0.05) or increased it moderately (0.05 < Δ*R*^2^ *≤* 0.2), with the exception of a low performance of CrAssphage for the sampling sites Munich and Augsburg and a low performance of PMMoV for Piding (Fig. 4). In the case of the sampling site Berchtesgaden, we observed a considerable increase (Δ*R*^2^ > 0.2) of the R^2^ value. Furthermore, we found that a normalization by PMMoV overall leads to higher R^2^ values than a normalization by CrAssphage.

Our findings indicate that the performance of surrogate viruses as normalization parameters in general and the difference between the performance of PMMoV and CrAssphage in particular are site-specific. This is in agreement with previous studies: Whereas Graham et al. ^33^ stated no substantial improvement when normalizing by surrogate viruses and Ai et al. ^17^ and Xie et al. ^34^ observed a deterioration of correlation with clinical data, Wilder et al. ^35^ found considerable positive correlations only when normalizing by CrAssphage. Further studies, comparing different sampling sites, reported improvements of the correlation for some locations but deteriorations for others ^18,24,45^. As for the comparison of the performance of PMMoV and CrAssphage, Nagarkar et al. ^26^ observed that PMMoV-normalization overall led to higher R^2^ values, whereas Greenwald et al. ^19^ reported the opposite. So far, the reasons behind the site-specific performance of surrogate viruses remain unclear ^18,26^.

For our results, a possible explanation for the overall good performance of PMMoV and CrAssphage is the low spatial and temporal variation of the surrogate virus biomarkers stated above (Fig. 1), as this indicates their suitability as normalization parameters ^19^. As for the lower performance of CrAssphage compared to PMMoV: this can be explained by the larger CV values observed for CrAssphage (Fig. 1), as a larger temporal variation of the normalization parameter blurs the dynamic of the SARS-CoV-2 biomarkers. This is further supported by the finding that for the only sampling site where CrAssphage had a lower CV value than PMMoV, Piding, normalization by CrAssphage led to a higher R^2^ value than by PMMoV. The considerable site-specific performance of surrogate viruses at the sampling site Berchtesgaden will be discussed in detail in the next section.

The best overall normalization performance was achieved with MFI: R^2^ values are generally either slightly or moderately increased, in the case of Berchtesgaden even considerably (Δ*R*^2^ = 0.49). The only decrease of R^2^ could be observed for Munich and is linked to the notable decrease of the R^2^ value for CrAssphage for the same sampling site. Purely mathematically speaking, the MFI therefore emerged as the best normalization parameter in our analysis. However, as both PMMoV and CrAssphage have to be determined by a PCR analysis in order to apply MFI, using this parameter is linked to higher expenses and does not lead to substantially better results than a normalization by CrAssphage or PMMoV alone. For datasets that comprise already data of several fecal indicators (not only CrAssphage and PMMoV, but also Bacteroides HF183 16S rRNA and human 18S rRNA, etc.), we nevertheless recommend testing the normalization by a mixed fecal indicator.

In recent publications, other normalization parameters than the ones considered here (PMMoV, CrAssphage, electrical conductivity, flow) were investigated. Informed by previous WBE studies ^46^, ammonia ^14,23,28^ and total nitrogen ^27^ were applied for normalization, but not compared to other parameters. Comparing the correlation between normalized SARS-CoV-2 wastewater data and clinical prevalence data, Xie et al. ^34^ found lower correlations for ammonia than for Acesulfame K; however, Pearson’s r in this study was generally moderate (max. 0.^6^1). In two studies, Bacteroides HF183 16S rRNA and human 18S rRNA were found to have higher temporal and spatial variation than PMMoV and CrAssphage and were discarded from further analysis ^18,19^. The performance of HF183 16S rRNA as a normalization parameter, just as with PMMoV and CrAssphage mentioned above, seems to be site-specific as it improved the correlations to clinical prevalence data for some sites, but deteriorated them for others ^24,26^.

#### 3.2.3 Dealing with a Complex Flow Situation: Sampling Site Berchtesgaden

The sampling site Berchtesgaden is a particular case that merits a detailed discussion. Compared to the other sampling sites, the coefficient of determination for the unnormalized data is particularly low (Fig. 4). The weak relationship between unnormalized SARS-CoV-2 data and clinical prevalence data can be explained by the particular flow situation in the service area of the WWTP Berchtesgaden. The high groundwater infiltration of up to 54% depends on the local groundwater level and thus, in a delayed and non-linear way, on the extent of previous precipitation events. Infiltrating groundwater then leads to a dynamic dilution of the wastewater that distorts the SARS-CoV-2 biomarker concentration in wastewater.

The coefficients of determination for flow-normalized SARS-CoV-2 biomarker data for all sampling sites did not differ notably from the unnormalized data (Δ*R*^2^ *≤* ± 0.0^6^). For instance, at the sampling site Teisendorf, a linear regression between clinical data and unnormalized data leads to *R*^2^ = 0.70, for flow-normalized data to *R*^2^ = 0.74 (Fig. 4). This is similar for the sampling site Berchtesgaden: the particularly low coefficient of determination for unnormalized data (*R*^2^ = 0.24) is marginally altered to *R*^2^ = 0.22 for flow-normalized data (Fig. 4 and 5). An explanation for this could be that normalizing by flow, a parameter that shows a strong relationship with precipitation data (Fig. S4), is an adjustment of the data enabling to account for stormwater dilutions when unnormalized SARS-CoV-2 biomarker data already has a strong relationship with clinical prevalence data. In contrast, in the particular case of a weak relationship between SARS-CoV-2 biomarker data and clinical prevalence data at the sampling site Berchtesgaden, flow-normalization cannot considerably improve the coefficient of determination. A similar observation can be made for normalization by electrical conductivity (Fig. 4).

**Figure 5:**
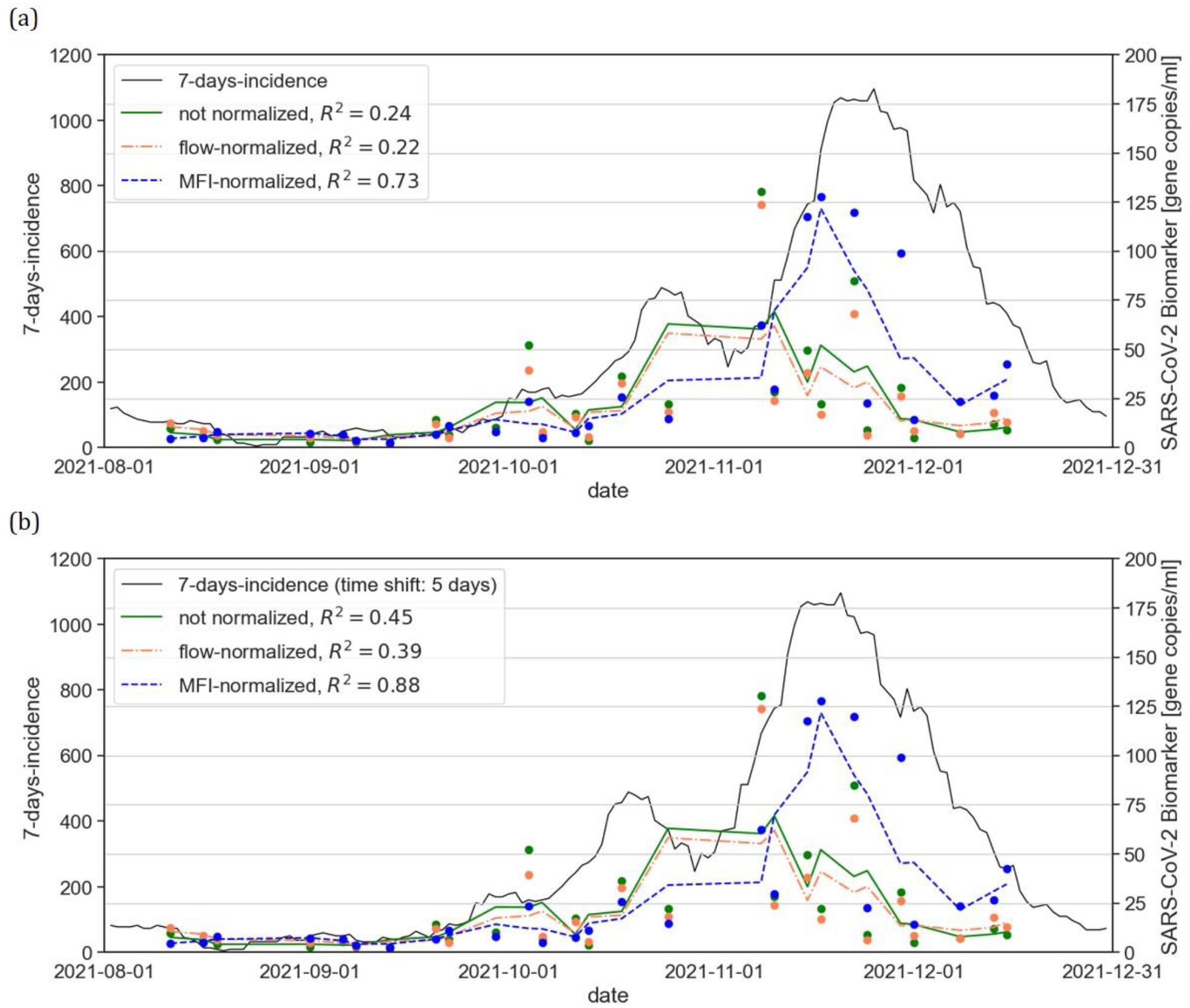
Timeline of 7-days-incidence and SARS-CoV-2 biomarker data with various normalizations for sampling site Berchtesgaden; without time shift of 7-days-incidence (a) and with time shift of 5 days (b).

In difference to this, normalizing by surrogate viruses shows promising results for the sampling site Berchtesgaden, improving the coefficient of determination considerably (Fig. 4 and 5). This can be explained by the particular nature of surrogate viruses as fecal markers: As PMMoV and CrAssphage biomarker concentrations are closely linked to human fecal shedding ^30,32^, they strongly depend on the fecal load in wastewater. For instance, it has been shown that PMMoV biomarker concentration increases with an increase of fecal contamination ^31^. Therefore, all parameters affecting the fecal load in wastewater, e.g. human shedding, stormwater dilution, but also dynamic groundwater infiltration, are accounted for by surrogate viruses because they directly mirror changes in fecal concentration. This detailed analysis suggests that surrogate-virus-normalization is especially interesting for sampling sites with high groundwater infiltration and possibly also for other particular flow situations, e.g. substantial and fluctuating industrial wastewater dischargers.

In previous investigations, lead times of the SARS-CoV-2 biomarker data ahead of the clinical data were stated ^7,13,16^. When accounting for this lead time for the sampling site Berchtesgaden, we observed a substantial improvement of the coefficients of determination for all normalizations for a time shift of 2 to 8 days with an optimum at 5 days (Fig. S6). Although improving the absolute R^2^ values for all normalizations, this time shift does not alter the relative differences: normalization by surrogate viruses continued to achieve considerably higher R^2^ values than unnormalized or flow-normalized data (Fig. 5 and S6). Accounting for the lead time of wastewater data ahead of clinical data was not the focus of the present study; however, our results indicate that this time-adjustment can further improve the relationship between normalized SARS-CoV-2 biomarker data and clinical prevalence data.

## 4. Conclusions

- Our analysis of the spatial variation of normalization parameters suggests that differences between sampling sites reflected characteristics of flow situation at a particular site, e.g., industrial wastewater dischargers or degree of groundwater infiltration. Knowing these characteristics helps to judge the suitability of the normalization parameter for a specific site.
- Surrogate viruses PMMoV and CrAssphage exhibited a low temporal variation and no noticeable seasonal trend, indicating their suitability as normalization parameters. CrAssphage showed generally higher temporal variation than PMMoV. Electrical conductivity and total daily flow followed seasonal trends, most likely because of the seasonal use of de-icing salt and the prevalent precipitation regime, respectively.
- Our findings indicate that the performance of surrogate viruses as well as flow and hydro-chemical data are site-specific. It is recommended to test the suitability of normalization parameters individually for specific sewage systems.
- Our results suggest that normalization by flow data reacts distinctly to precipitation events and adjusts SARS-CoV-2 biomarker concentrations accordingly so that relevant changes in the infection pattern can be observed also during precipitation periods. However, coefficients of determination between flow-normalized SARS-CoV-2 biomarker data and clinical prevalence data are generally lower than for surrogate-virus-normalized data.
- The data from the sampling at the WWTP Berchtesgaden, a site suffering from severe groundwater infiltration, suggests that weak relationships between unnormalized SARS-CoV-2 biomarker data and clinical prevalence data can be considerably improved by normalization with surrogate viruses, but not with flow or electrical conductivity data. Adjusting for the lead time of wastewater data ahead of clinical prevalence data further improved this relationship.
- Normalization using PMMoV led generally to higher R^2^ than CrAssphage-normalization, most likely due to the lower temporal variation observed in our data. The best overall normalization performance was achieved with the mixed fecal indicator based on PMMoV and CrAssphage; nevertheless, the improvements of the R^2^ values are not notably higher than with PMMoV or CrAssphage alone. For further studies, we recommend to evaluate temporal and financial expenditures for measuring both surrogate viruses.

## Data Availability

All data produced in the present work are contained in the manuscript. Original raw data are available upon reasonable request to the authors

## Acknowledgements

This study was financially supported by the German Federal Ministry of Education and Research as part of the funding program Sustainable Water Management (NaWaM-RiSKWa) (Biomarker, grant number 02WRS1557). We thank the staff of the WWTPs Augsburg, Berchtesgaden, Freilassing, Karlsruhe, München, Piding, and Teisendorf as well as the health department of the county of Berchtesgadener Land for their support. We also thank Marion Woermann, Heidrun Mayrhofer, Lucia Maciossek, Meenakshi Prasad, and Mohammad Sheryaar Khan from TUM as well as Rabea Suhrborg, Carmen Kraffert, and Marie Weihnacht from TZW for their assistance with laboratory work.

## Supporting Information

Details of materials and methods (Supplemental Text 1); One-Step-RT-PCR program (Table S1); spatial and temporal variation of daily flow per capita and electrical conductivity (Figure S1); timeline of daily flow data, conductivity data, and precipitation data (Figures S2 and S3); coefficients of determination of linear regression models for precipitation data and normalization parameters (Figure S4); timeline of precipitation data, SARS-CoV-2 biomarker data, and 7-days-incidence (Figure S5); coefficients of determination of linear regression models for clinical prevalence data and SARS-CoV-2 biomarker data with different time shifts (Figure S6) (DOC)

