## Supplemental Material for "Wastewater-Based Epidemiology for SARS-CoV-2 Biomarkers: Evaluation of Normalization Methods in Small and Large Communities in Southern Germany"

### Supplemental Information

#### List of Supplemental Text, Tables, and Figures

**Supplemental Text 1** Details of materials and methods

**Table S1** One-Step-RT-PCR program

**Figure S1** Spatial and temporal variation of daily flow per capita and electrical conductivity for different sampling sites

**Figure S2** Timeline of daily flow data and precipitation data a different sampling sites

**Figure S3** Timeline of conductivity data and precipitation data at different sampling sites

**Figure S4** Coefficients of determination of linear regression models for precipitation data and various normalization parameters

**Figure S5** Timeline of precipitation data, SARS-CoV-2 biomarker data (unnormalized and flow-normalized) and 7-days-incidence for different sampling sites

**Figure S6** $R^2$ of linear regression models for clinical prevalence data and SARS-CoV-2 biomarker data (various normalizations) for the sampling site Berchtesgaden with different time shifts

### Supplemental Text 1: Details of Materials and Methods

### Sample Processing

Samples from Munich and Augsburg were processed at the Technical University of Munich, Chair of Urban Water Systems Engineering in Garching, Germany according to the following protocol: After maximum 48 h of storage at 4 °C, the concentration and nucleic acid extraction of sewage samples were performed with the *Wizard Enviro Total Nucleic Acid Kit* (Promega), following the manufacturer's recommendations. Extracts were eluted in 40-50 µl nuclease-free water and stored at -80 °C. Subsequently, digital Polymerase Chain Reaction (dPCR) was performed on the wastewater extracts. Three SARS-CoV-2 specific gene fragments were quantified: the nucleocapsid gene (N2), envelope gene (E), and a sequence from the open reading frame region (ORF). Additionally, the fecal marker PMMoV was qualitatively determined for internal process control with each dPCR assay. Primer sequences can be found in Ho et al. [1]. The dPCR analysis was conducted on the *QIAcuity One Digital PCR System* (Qiagen) using the *QIAcuity One-Step Viral RT-PCR Kit* (Qiagen). This dPCR Kit was used as described by the manufacturer but with the supplement of 1.25 % GoScript Reverse Transcriptase (Promega) to the total reaction volume to alleviate inhibition of the reverse transcription by matrix effects. A negative control (nuclease-free water (Qiagen)) and two positive controls (PMMoV (PMMoV RNA, $4 \times 10^6$ copies/µl, Promega) and synthetic SARS-CoV-2 (SARS-CoV-2 (N+E) RNA, $4 \times 10^6$ copies/µl, Promega)) were included in each dPCR assay. The imaging and PCR cycling was done in a *QIAcuity Nanoplate 26k* 24-well plate (Qiagen) using 4 µl of template with the One-Step-RT-PCR program described in Table S1. The *QIAcuity Software Suite* (Qiagen) was used to analyze the raw data, which were back-calculated as gene copies per ml of wastewater based on the volume of the original wastewater sample. A detection limit (LOD) of 1 gene copy/ml wastewater was determined.

Samples from the WWTP Karlsruhe and the four small communities (Berchtesgaden, Freilassing, Piding, Teisendorf) were processed at the *TZW: DVGW-Technologiezentrum Wasser, Department of Water Microbiology* in Karlsruhe, Germany as described by Ho et al. [1]. Three SARS-CoV-2 specific sequences were quantified: E, ORF and a sequence from the polymerase region (RdRp). For this process, an LOD of 2.5 gene copies/ml was determined. The concentrations of biomarkers of the fecal markers PMMoV and CrAssphage for samples from all sites were also determined in this lab according to the same dPCR protocol.

**Linear regression analysis with precipitation as explanatory variable**

Linear regression analysis with the daily sum of precipitation $[mm/d]$ as explanatory variable was performed with a regression model not forced through zero (Statsmodels v0.12.2) (Fig. S4). The coefficients of determination $R^2$ were classified as weak ($R^2 \leq 0.3$), moderate ($0.3 < R^2 \leq 0.5$), and strong ($R^2 > 0.5$) for the linear regression of precipitation data and normalization parameters. This analysis was performed for the precipitation data of the sampling day, the previous day, as well as the sum of the previous and the sampling day as the flow times for the sampling sites differ and can reach up to 24h for large sewersheds. Days without precipitation were excluded from this analysis.

**Data Processing Workflow for SARS-CoV-2 Biomarker Concentrations**

The SARS-CoV-2 biomarker concentrations from the PCR analysis were analyzed using an automated workflow. The first step was the application of PMMoV and CrAssphage abundances as internal process control to account for errors during sampling, transport, or sample processing. Samples with $log_{10}$ gene copies/ml values of one of the surrogate viruses outside of the interval of $\mu \pm 2\ \sigma$ were discarded. In total, 10 sampling values were excluded from further analysis due to this filter.

In a next step, values of SARS-CoV-2 biomarker concentrations were separated in positive findings and values below the LOD. If less than two of the three biomarker genes determined had values above the LOD, the average value of that sampling day was reported as half of the LOD (1.25 resp. 0.5 gene copies/l). If at least two biomarker genes were above the LOD, the average value of biomarkers was computed as the mean of these gene values (biomarker genes with values reported below the LOD were assigned the numerical value of half the LOD). In total, 27 samples were classified as below the LOD. These were mainly from the beginning of August 2021 from the small sampling sites, corresponding to a period of few clinical cases. Subsequently, additional information on the flow situation during sampling, provided by the WWTPs, was used to exclude data points from further analysis. Based on this filter, the samples from Teisendorf on 2021-08-30 and 2021-09-01 were excluded because the WWTP reported a major combined sewer overflow due to a heavy precipitation event for these sampling days. In a final step, the rolling average over 3 sampling days (corresponding to 7 days) was computed for the SARS-CoV-2 average genes in order to smooth the data without masking short-term trends.

### Application of Normalization Parameters on SARS-CoV-2 Biomarker Data

Normalizing by most parameters (CrAssphage, PMMoV, MFI, electrical conductivity) means dividing by them according to the following formula (1):

$$\text{Genes}_{avg,norm,P} = \text{Genes}_{avg} \cdot \frac{\overline{P_{dry}}}{\text{P}} \qquad (1)$$

In contrast, flow represents the volume of the wastewater; normalization by flow values can therefore be considered a conversion to SARS-CoV-2 biomarker loads in wastewater and is computed as follows (2):

$$\mathrm{Genes}_{avg,norm,Q} = \mathrm{Genes}_{avg} \cdot \frac{\mathrm{Q}}{\overline{Q_{dry}}} \qquad (2)$$

$\overline{P_{dry}}$ and $\overline{Q_{dry}}$ are the mean values of the normalization parameters on dry days and are added to the normalization procedure in order to keep the unit [gene copies/ml] and the order of magnitude of $\mathrm{Genes}_{avg,norm}$ and $\mathrm{Genes}_{avg}$ consistent. This assured that different normalization methods could be compared graphically in the same figure.

**Table S1**: One-Step-RT-PCR program.

| Step | Number of cycles | Time | Temperature |
|---|---|---|---|
| Reverse Transcription | 1 cycle | 40 min | 50 °C |
| PCR initial heat activation | 1 cycle | 2 min | 95 °C |
| Denaturation | 2-step cycling (40 cycles) | 5 s | 95 °C |
| Combined annealing / extension | | 60 s | 60 °C |

**Figure S1**: Spatial and temporal variation of daily flow per capita (a) and electrical conductivity (b) for different sampling sites.

(a)

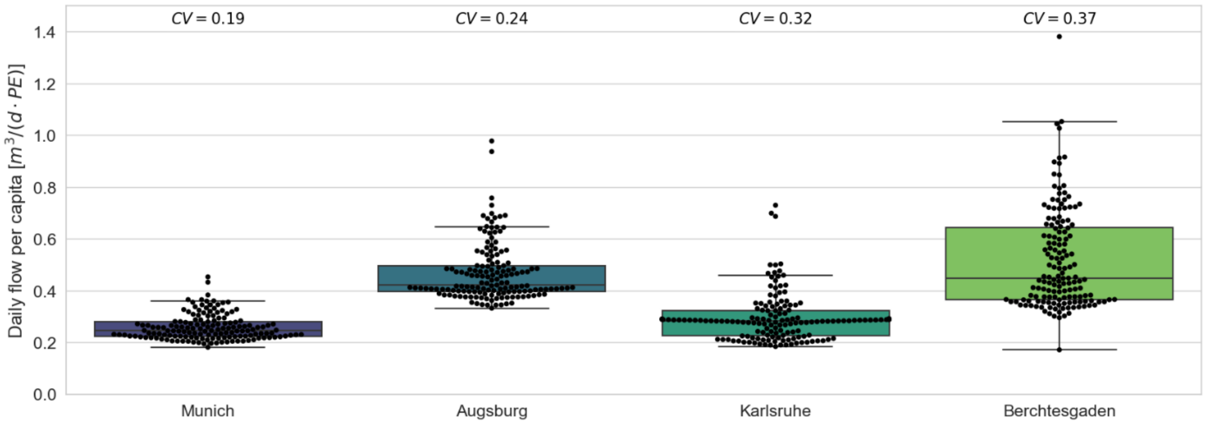


(b)

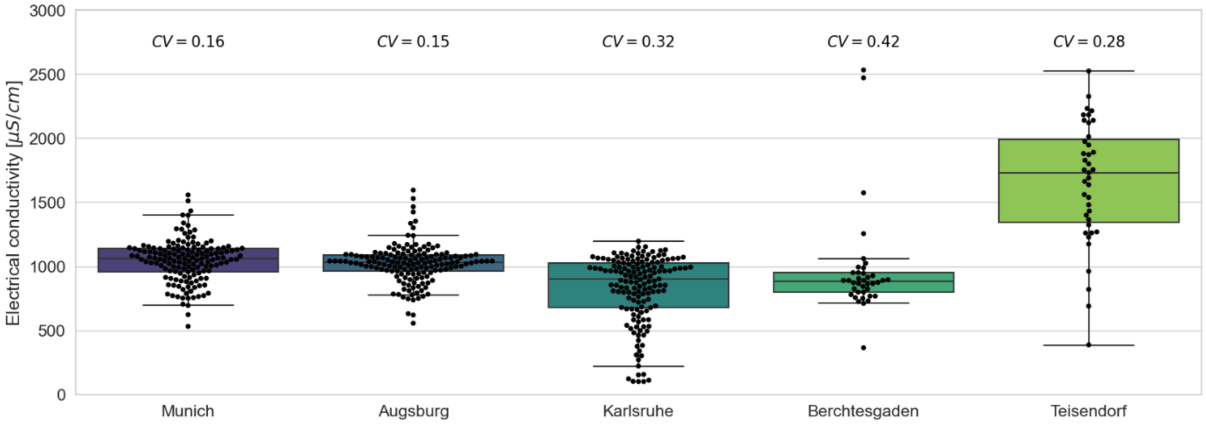

**Figure S2**: Timeline of daily flow data and precipitation data at sampling sites Munich (a), Karlsruhe (b), and Berchtesgaden (c).

(a)

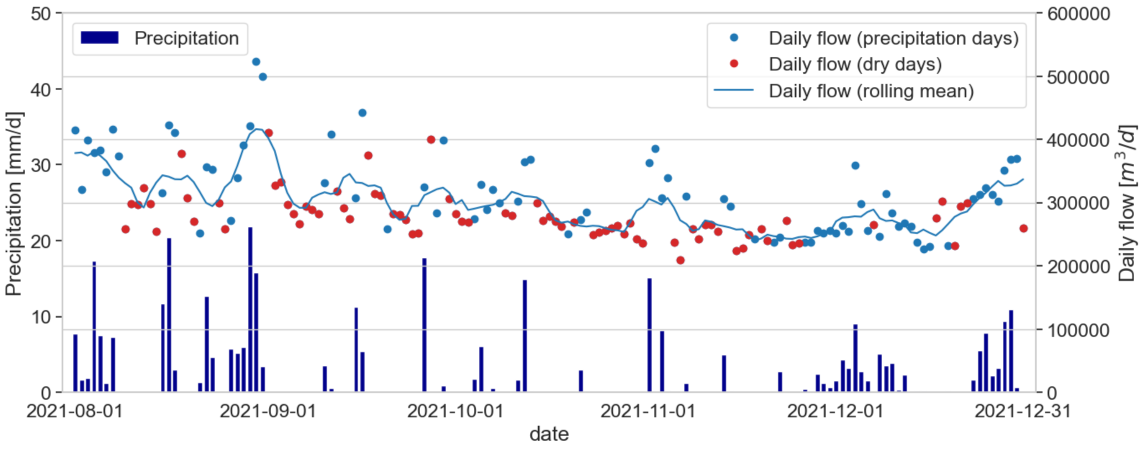


(b)

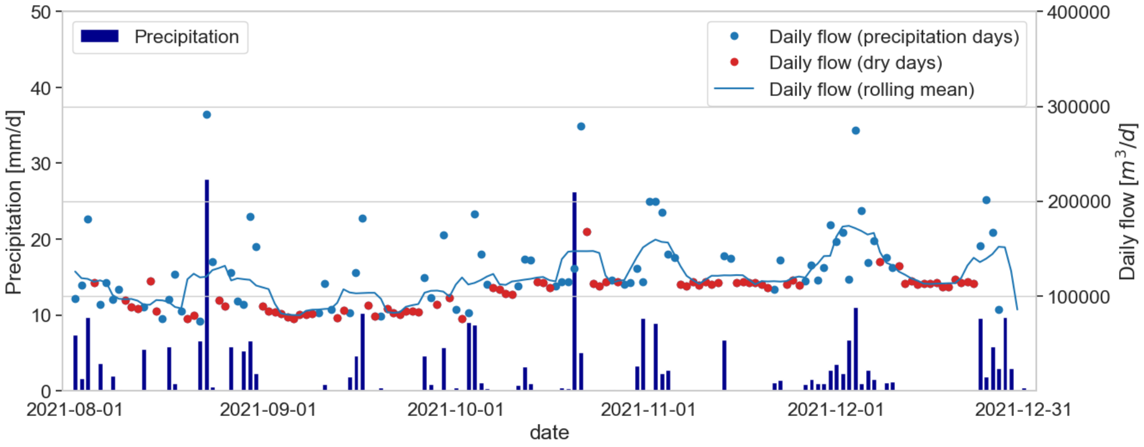


(c)

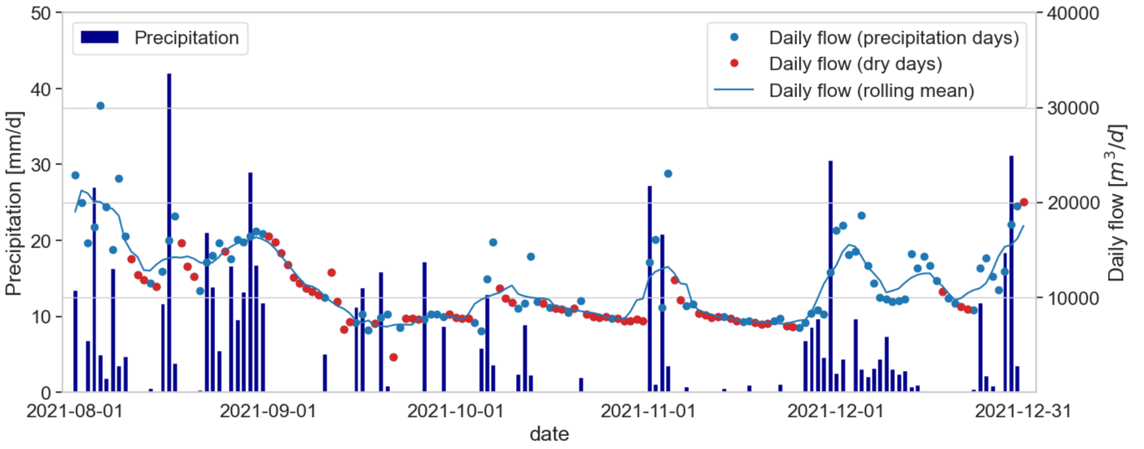

**Figure S3**: Timeline of conductivity data and precipitation data at sampling sites Augsburg (a), Munich (b), Karlsruhe (c), Berchtesgaden (d), and Teisendorf (e)

(a)

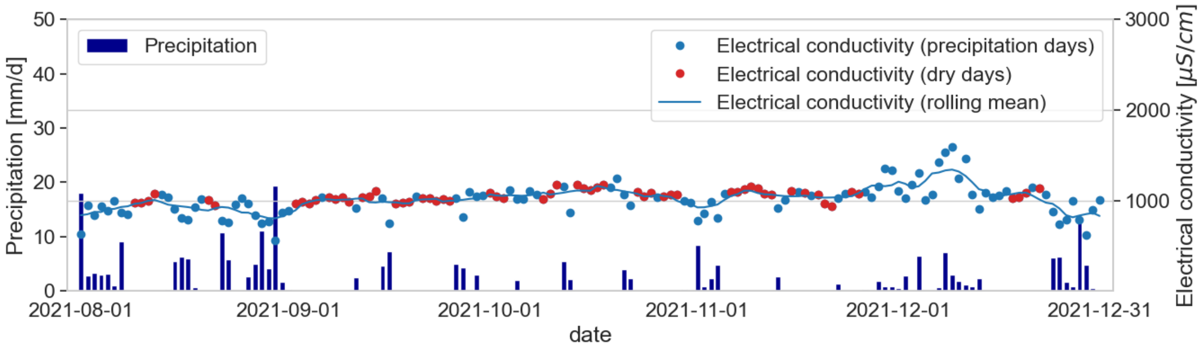


(b)

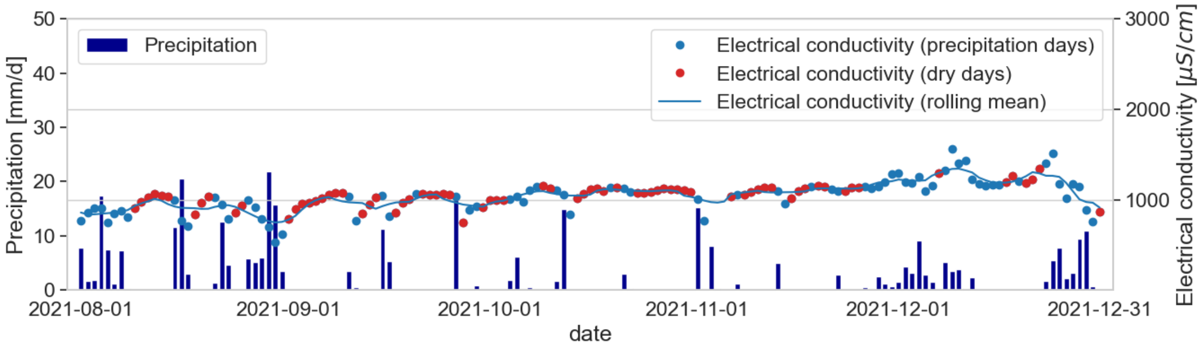


(c)

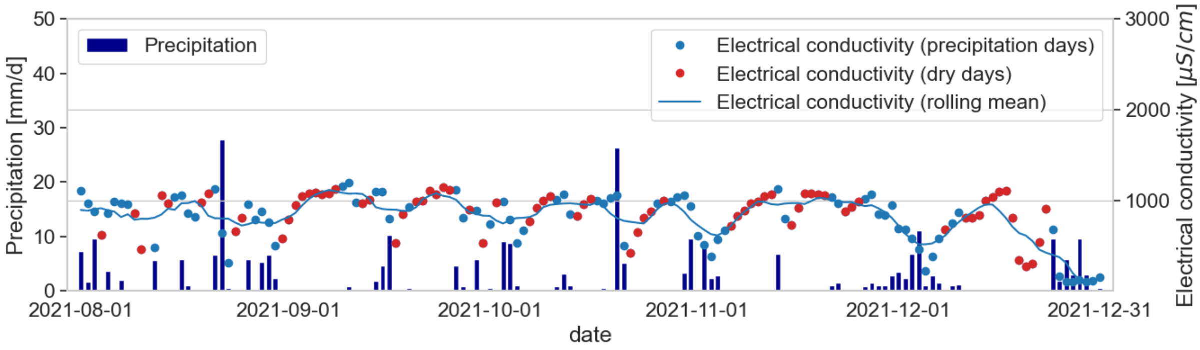


(d)

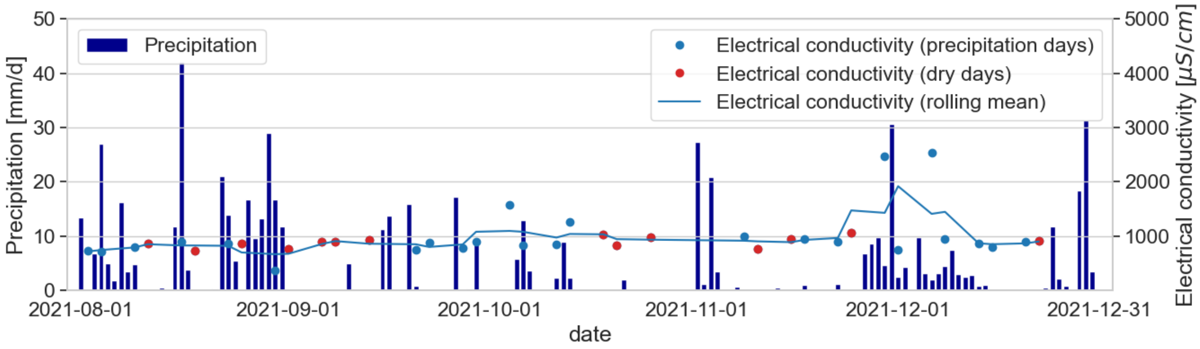
Precipitation
Electrical conductivity (precipitation days)
Electrical conductivity (dry days)
Electrical conductivity (rolling mean)
Precipitation [mm/d]
Electrical conductivity [μS/cm]
date
2021-08-01
2021-09-01
2021-10-01
2021-11-01
2021-12-01
2021-12-31

(e)

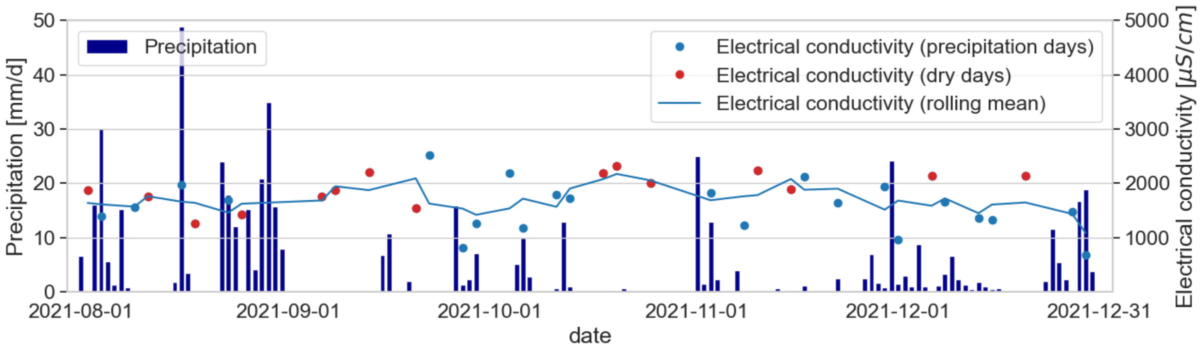
Precipitation
Electrical conductivity (precipitation days)
Electrical conductivity (dry days)
Electrical conductivity (rolling mean)
Precipitation [mm/d]
Electrical conductivity [μS/cm]
date
2021-08-01
2021-09-01
2021-10-01
2021-11-01
2021-12-01
2021-12-31

**Figure S4**: (a) Linear regression model for precipitation data of current day and flow in Augsburg; (b-d) $R^2$ of linear regression for precipitation data of current day (b), previous day (c), and sum of previous and current day (d) for various normalization parameters and sampling sites. The coefficients of determination $R^2$ were classified as weak ($R^2 \leq 0.3$, red), moderate ($0.3 < R^2 \leq 0.5$, yellow), and strong ($R^2 > 0.5$, green).

(a)

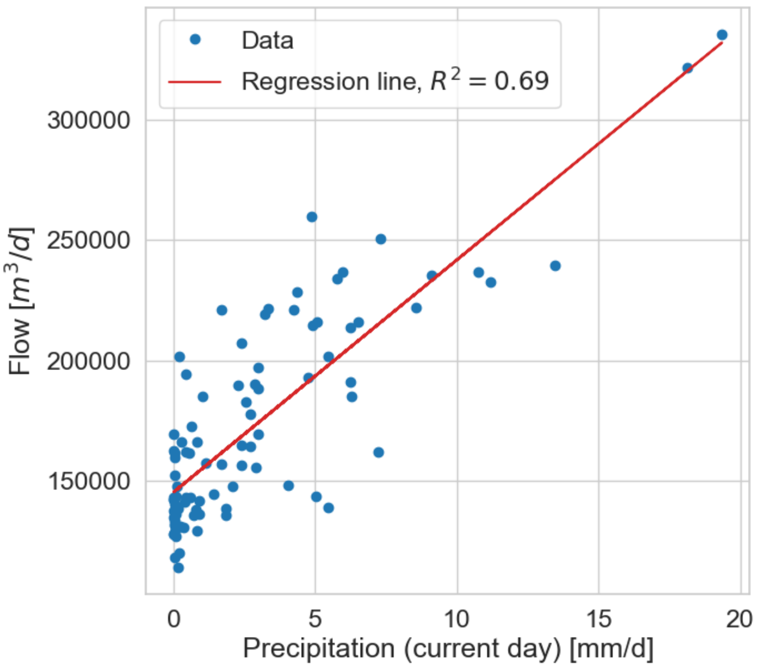


(b)

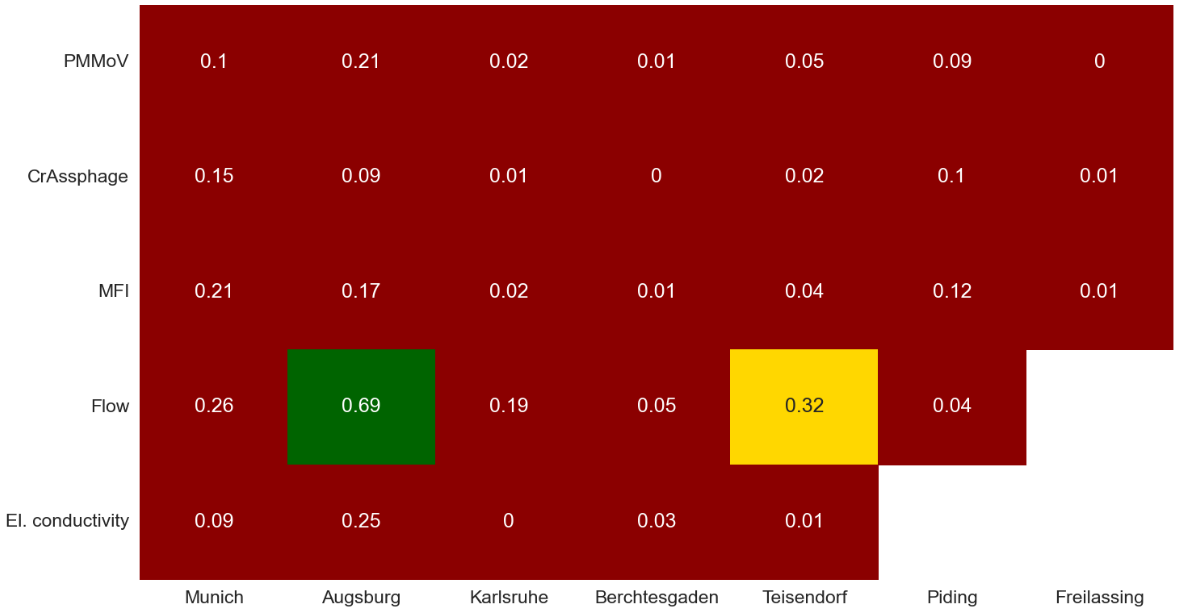


(c)

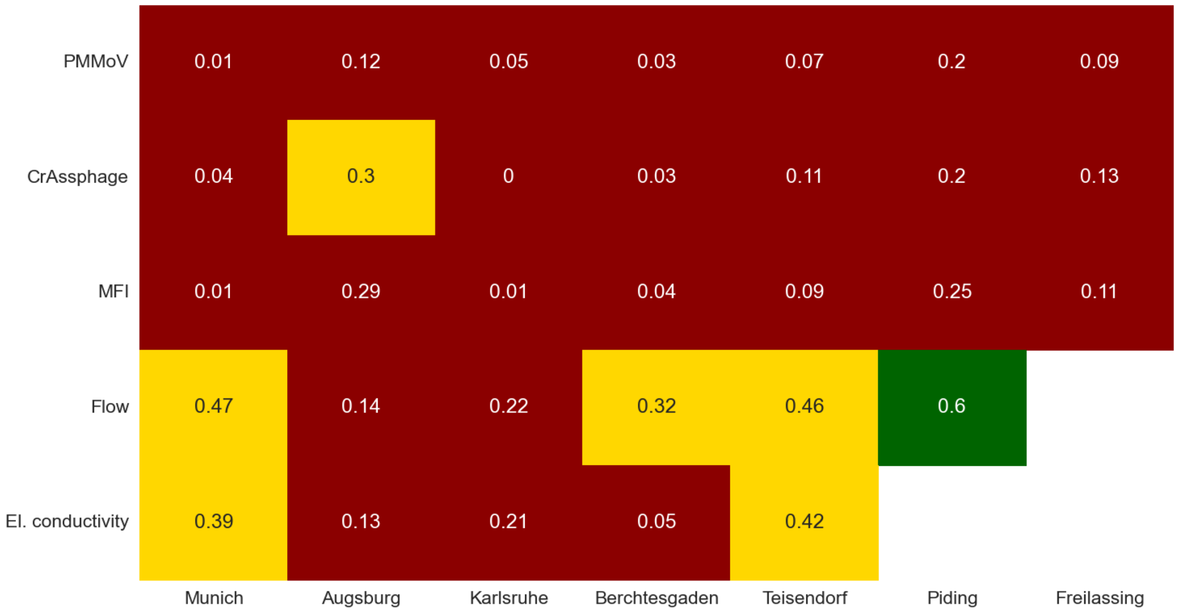
PMMoV
0.01
0.12
0.05
0.03
0.07
0.2
0.09
CrAssphage
0.04
0.3
0
0.03
0.11
0.2
0.13
MFI
0.01
0.29
0.01
0.04
0.09
0.25
0.11
Flow
0.47
0.14
0.22
0.32
0.46
0.6
El. conductivity
0.39
0.13
0.21
0.05
0.42
Munich
Augsburg
Karlsruhe
Berchtesgaden
Teisendorf
Piding
Freilassing

(d)

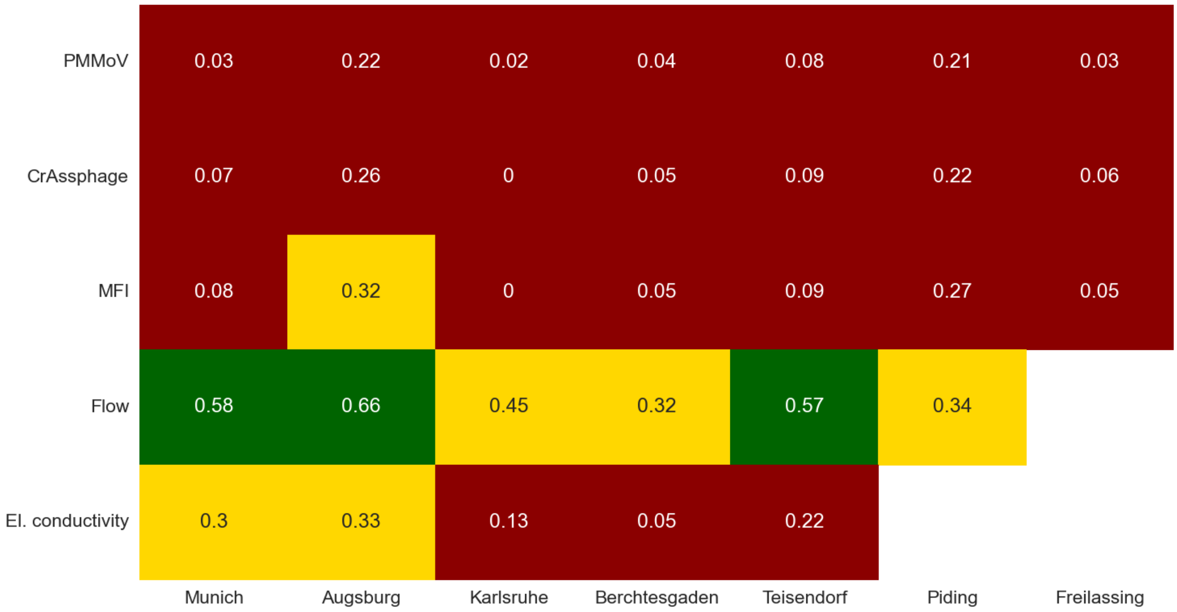
PMMoV
0.03
0.22
0.02
0.04
0.08
0.21
0.03
CrAssphage
0.07
0.26
0
0.05
0.09
0.22
0.06
MFI
0.08
0.32
0
0.05
0.09
0.27
0.05
Flow
0.58
0.66
0.45
0.32
0.57
0.34
El. conductivity
0.3
0.33
0.13
0.05
0.22
Munich
Augsburg
Karlsruhe
Berchtesgaden
Teisendorf
Piding
Freilassing

**Figure S5**: Timeline of precipitation data, SARS-CoV-2 biomarker data (unnormalized and flow-normalized) and 7-days-incidence for sampling sites Munich (a), Karlsruhe (b), Piding (c), and Teisendorf (d) in late summer and early fall 2021.

(a)

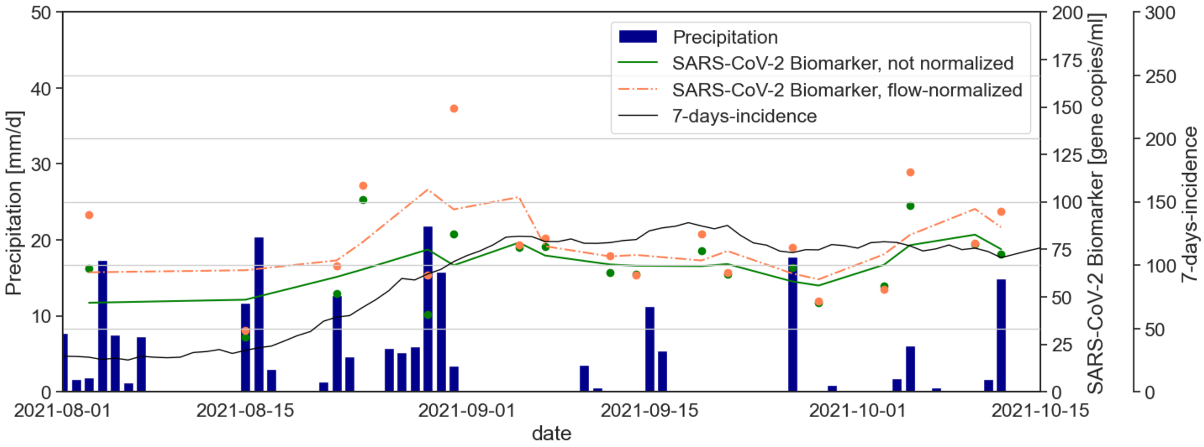


(b)

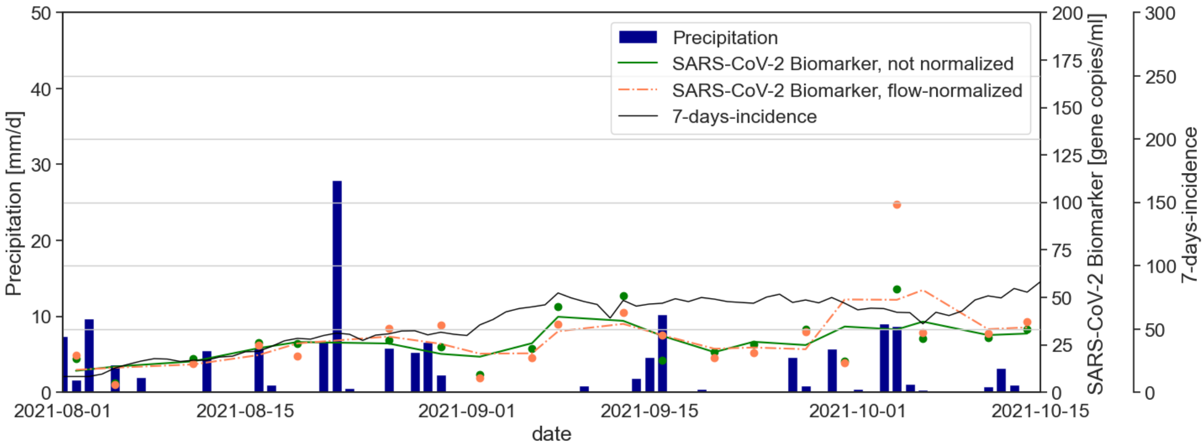

(c)

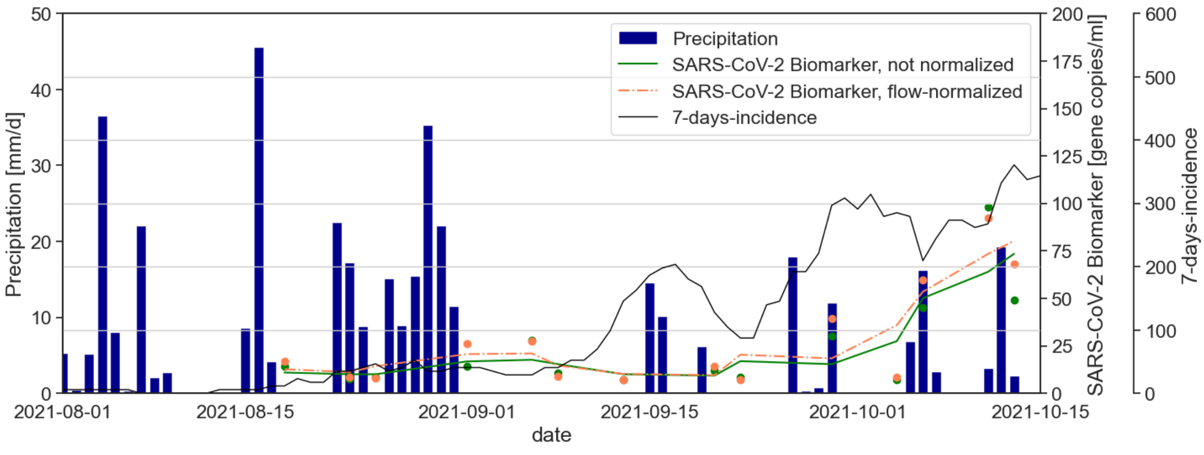


(d)

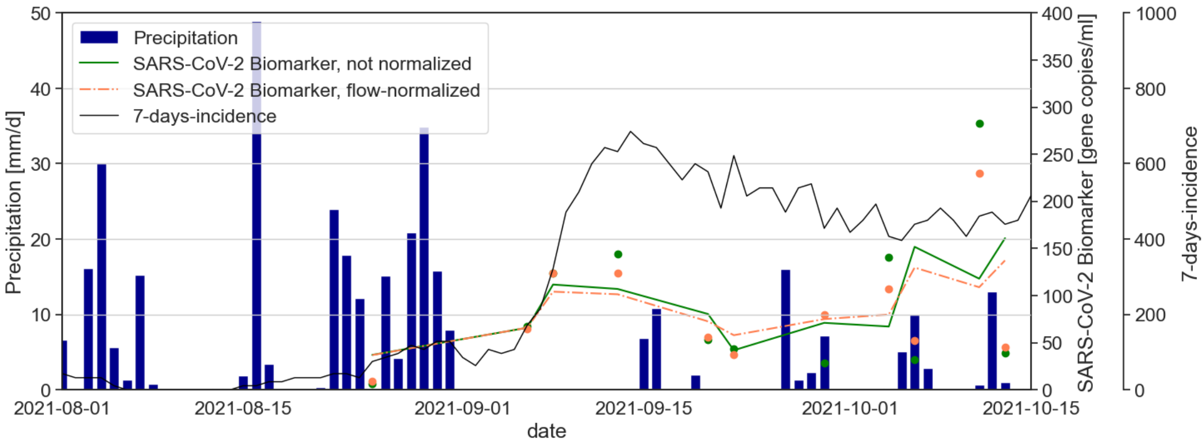


**Figure S6**: $R^2$ of linear regression for clinical prevalence data and SARS-CoV-2 biomarker data (various normalizations) for the sampling site Berchtesgaden with different time shifts from 0 to 15 days for the clinical prevalence data.

| | 0 | 1 | 2 | 3 | 4 | 5 | 6 | 7 | 8 | 9 | 10 | 11 | 12 | 13 | 14 | 15 |
|---|---|---|---|---|---|---|---|---|---|---|---|---|---|---|---|---|
| Unnormalized | 0.24 | 0.27 | 0.36 | 0.38 | 0.4 | 0.45 | 0.43 | 0.5 | 0.53 | 0.56 | 0.6 | 0.63 | 0.64 | 0.69 | 0.69 | 0.71 |
| CrAssphage | 0.68 | 0.72 | 0.8 | 0.84 | 0.83 | 0.83 | 0.8 | 0.8 | 0.8 | 0.71 | 0.67 | 0.63 | 0.6 | 0.6 | 0.54 | 0.52 |
| PMMoV | 0.72 | 0.77 | 0.86 | 0.87 | 0.87 | 0.88 | 0.85 | 0.86 | 0.85 | 0.78 | 0.77 | 0.74 | 0.72 | 0.74 | 0.68 | 0.66 |
| MFI | 0.73 | 0.78 | 0.87 | 0.88 | 0.88 | 0.88 | 0.85 | 0.85 | 0.84 | 0.75 | 0.73 | 0.69 | 0.67 | 0.67 | 0.61 | 0.58 |
| Flow | 0.22 | 0.23 | 0.31 | 0.32 | 0.35 | 0.39 | 0.37 | 0.44 | 0.47 | 0.49 | 0.54 | 0.57 | 0.58 | 0.62 | 0.63 | 0.65 |
| El. conductivity | 0.22 | 0.25 | 0.34 | 0.37 | 0.39 | 0.44 | 0.42 | 0.5 | 0.54 | 0.56 | 0.61 | 0.65 | 0.66 | 0.7 | 0.7 | 0.72 |
